# Systematic Evaluation of Common Natural Language Processing Techniques to Codify Clinical Notes

**DOI:** 10.1101/2022.10.10.22280852

**Authors:** Nazgol Tavabi, Mallika Singh, James Pruneski, Ata M. Kiapour

## Abstract

Proper codification of medical diagnoses and procedures is essential for optimized health care management, quality improvement, research, and reimbursement tasks within large healthcare systems. Assignment of diagnostic or procedure codes is a tedious manual process, often prone to human error. Natural Language Processing (NLP) have been suggested to facilitate these manual codification process. Yet, little is known on best practices to utilize NLP for such applications. Here we comprehensively assessed the performance of common NLP techniques to predict current procedural terminology (CPT) from operative notes. CPT codes are commonly used to track surgical procedures and interventions and are the primary means for reimbursement. The direct links between operative notes and CPT codes makes them a perfect vehicle to test the feasibility and performance of NLP for clinical codification. Our analysis of 100 most common musculoskeletal CPT codes suggest that traditional approaches (i.e., TF-IDF) can outperform resource intensive approaches like BERT, in addition to providing interpretability which can be very helpful and even crucial in the clinical domain. We also proposed a complexity measure to quantify the complexity of a classification task and how this measure could influence the effect of dataset size on model’s performance. Finally, we provide preliminary evidence that NLP can help minimize the codification error, including mislabeling due to human error.

## Introduction

Advancements in natural language processing (NLP) have led to increased interest in health care research and quality improvement. The development of automated pipelines to process large volumes of clinical notes can optimize healthcare operation (e.g., quality improvement, resource management, billing) and improve clinical care (e.g., evidenced-based clinical decision making). As up to 80% of the electronic medical record is comprised of this unstructured documentation [1], [2], NLP is postured to become an increasingly valuable tool for processing this data. Currently, a significant challenge in this process is the identification and development of proper techniques that can effectively characterize unstructured clinical notes (e.g., text representation) and make predictions using that information to improve efficiency and provide high-value patient care.

Over the past decade, several NLP techniques have been developed and adapted to process clinical notes, ranging from simple and traditional approaches such as bag of words to more computationally expensive and complex approaches such as transformers and large language models. Yet, there is a paucity of studies to systematically evaluate their relative performance in clinical domain. Codification of diagnoses (ICD, International Classification of Diseases) or procedures (CPT, Current Procedural Terminology) from clinical and procedure notes offer a unique opportunity to systematically assess the performance of NLP techniques in clinical domains. Efficient and accurate coding of clinical notes (e.g., billing codes or diagnostic codes) impacts the entire healthcare industry, including healthcare systems, care givers, insurance companies, and policy makers. The practice of assigning codes is a complex and labor-intensive process that is prone to human error. Failure to code correctly can result in inadequate patient care and may lead to an increase in expenses or delays in the reimbursement process. Recent studies regarding the automation of the coding process have tried an array of techniques, ranging from traditional text matching to deep learning-based approaches, to categorize clinical notes [3]– [11].

The purpose of the current study is to systematically evaluate the ability of commonly used NLP techniques to predict CPT codes from unstructured operative notes. Operative notes are a specific subgroup of clinical notes that contain details of surgical procedures performed on patients and the primary source for CPT billing code assignment. Information in these operative notes is used in tasks such as cohort identification [12], quality assessment [13], and surgical procedures information extraction [14]–[16]. The clinical importance of operative notes along with their one-to-one relationship with CPT codes offer a unique opportunity to evaluate NLP performance in clinical domain. Considering the importance and resource intensity of manual CPT coding, an accurate scalable CPT prediction platform would help to optimize healthcare operations and reduce billing and reimbursement errors, and processing times. In this study, we compared the performance of three common NLP techniques (Term Frequency-Inverse Document Frequency (TF-IDF) [17], Doc2Vec [18], and Bidirectional Encoder Representations from Transformers (BERT) [19] to predict the 100 most common musculoskeletal CPT codes in a high-volume multi-site academic pediatric and young adult orthopedic surgery and sports medicine clinic. We further studied text-related factors influencing the model performance (i.e., complexity of CPTs). We hypothesized that NLP models could predict CPT codes with near-human accuracy, and that the state-of-the-art embedding-based models (e.g., BERT) would outperform traditional NLP approaches (e.g., TF-IDF) in predicting the CPT codes from the operative notes. Secondarily, we hypothesized that increased CPT complexity would be associated with decreased model performance.

## Method

### Data

Following IRB approval (IRB-P00037878), operative notes from patients with at least one encounter at any of the six Boston’s Children Hospital Orthopedic Surgery and Sports Medicine clinics from 2010 – 2020 were acquired (n=126,789 documents). The operative notes were filtered to include only procedures on the musculoskeletal system (CPT codes 20100 - 29999), resulting in 44,002 notes. The 100 most prevalent CPT codes in our dataset (Supplementary Table S1) were used as labels for the classification task.

### TF-IDF

TF-IDF is a statistical method for extracting features, or keywords, from text [17]. With this method, documents are represented as TF-IDF vectors where each feature (dimension) in the vector space corresponds to a unique word in the dataset. The weight of each term is proportional to the number of times it is repeated in the document. Additionally, terms are inversely weighted by the number of documents in the corpus that contain them (IDF). IDF helps to adjust for common words that do not carry much definitive value. The number of features in this representation is equal to the number of terms in the vocabulary, which is often quite large and can lead to overfitting and slow learning [20]. One common approach to minimize that risk is to conduct feature selection on TF-IDF vectors before training, which involves training the model only on a subset of features based on their relevance to the target variable. However, the proper feature size (K) selection is key in minimizing the risk of overfitting due to a large number of features or underfitting due to a lack of sufficient information. To address this important issue in the context of clinical note labeling, we evaluated the effect of feature space size on model performance. This was done by training and testing classifiers to predict CPT code from operative notes with changing TF-IDF feature space size (K). By applying TF-IDF to the dataset, around 42,000 words were identified, corresponding to 42,000 features. The following four classifications were completed using this method:

1. Classification based on all extracted features
2. Classification based on the top 100 most relevant keywords to the target CPT (identified using F-values from ANOVA test)
3. Classification based on top 500 most relevant keywords to the target CPT
4. Classification based on top K most relevant keywords, where K is a hyper-parameter that is optimized for each CPT to get the best performance.

For TF-IDF with variable feature size K, the search space for K was set as log-uniform so that the model has a higher chance of assigning a lower number to K to avoid overfitting.

### Doc2vec

Doc2vec [18] is a generalization of Word2vec [21], which is a technique used to represent words (tokens) as vector embeddings (points in a high-dimensional space) such that similar words have representations that are closer to each other. Doc2Vec uses the word embeddings generated by word2vec to vectorize the entire documents. Word2vec and Doc2vec use shallow networks to learn the context of words and sentences. Word2vec model is trained based on one of two approaches; either the model learns the embeddings of tokens such that it can be used to predict their context (surrounding words), or the model is trained such that the context (surrounding words) can predict the token. Similarly, Doc2vec model is trained such that either the model learns sentence/paragraph embeddings to predict a probability distribution of words in a sentence (context) given a randomly sampled word from that sentence (Distributed Bag of Words (DBOW) approach), or the model learns the embeddings such that it can predict a word given the sentence representation in addition to its surrounding words (Distributed Memory (DM) approach). To train Doc2vec representations, the following hyper-parameters were set:

- D: dimension of the embeddings
- W: window size (the W surrounding words which is referred to as context in the description of Doc2vec.)
- DM versus DBOW: Optimization approach, using either Distributed Memory or Distributed Bag of Words

### BERT

BERT is Start-Of-The-Art NLP model developed by Google which has been extensively used in multiple text processing domains and NLP tasks [19]. BERT is a deep neural network with multiple layers of encoders with bidirectional self-attention heads. Because of its many layers, BERT models require a large amount of training data. Hence, in most cases the pre-trained versions of these models are used. Furthermore, researchers have pre-trained the BERT models on more specific datasets, as to improve the model’s performance for specific tasks. In this study we use Clinical Bio-BERT [22], which is fine-tuned on medical literature (PubMed corpus) [23], and further fine-tuned on publicly available MIMIC clinical notes [24]. It’s worth noting that while BERT models can process sequences with max length of 512 tokens, clinical notes are usually longer. One way of dealing with this problem is to truncate the note and only feed in the first 510 tokens (2 tokens are reserved for special tokens in BERT), however since the details of the surgical procedure, required for proper prediction of the CPT code, are spread across the note this is not a viable solution for this task. To make sure all the information in the text are captured, notes were broken down to sub-sequences of less than 512 tokens, and their embeddings were extracted from BERT and then separately given to a classifier. This approach is the same as freezing the BERT model and only optimizing the classifier for the classification, which is a common approach for clinical note classification [25].

A challenge with this approach is getting note level embeddings based on word (token) level embeddings. Several approaches have been previously suggested, including averaging the embeddings of all words in the document, taking the maximum word embedding in the document as a way to select the most important features [26], or using the embeddings of a special token in the beginning of each sentence called the [CLS] token as a general representation of input sentence [27], [28]. Usually in a supervised setting the embedding of the [CLS] token is used. However, in an unsupervised setting (which is the case here), the [CLS] token does not carry much information. Here we generated the whole note representation using average and maximums of word embeddings and the CLS tokens in the clinical note and then compared the performance of each approach in predicting the CPT code using the following configurations:

- A_W: Average token/Word Embeddings
- M_W: Max of token/Word Embeddings
- A_CLS: Average of [CLS] embeddings
- M_CLS: Max of [CLS] embeddings
- X + Y: Concatenate embeddings of X and Y

### Classification

Support vector machine (SVM) with RBF kernel [29] was used as the classifier for all classifications. The generated features from one of the aforementioned approaches (i.e., TF-IDF, Doc2vec, and BERT) were used as input to the classifier to predict the CPT codes. Separate classifiers were trained for each CPT. Using the same classifier for all approaches enabled us to isolate the effect of the feature extraction approach on model performance. Model hyper-parameters (*C* & *γ* and *K* for TF-IDF with feature selection) were optimized with Hyperopt [30], which is a Python library for parameter tuning based on Bayesian optimization. The results of past evaluations are used to learn a probabilistic model for the objective function (i.e., minimizing the loss), and to choose the next set of hyper-parameter values based on that function. 20% of the data (by preserving the percentage of samples for each class) was used as test set and the training data was split into 5-fold cross-validation for fine-tuning. After choosing the best hyper-parameters, the model was trained on the whole train data and evaluated on test data.

### Model Performance Evaluation

We assessed the model’s performance in predicting the CPT codes with the area under the receiver operating characteristic curve (AUROC). We also calculated the accuracy, sensitivity, and specificity across all predictions. We first compared the performance of classifiers within each feature extraction approach (i.e., TF-IDF, Doc2vec, and BERT). We then compared the best-performing models from each approach using a Critical Difference Diagram [31], [32]. The Critical Difference Diagram is an approach to compare multiple classifiers over multiple data sets, and it ranks the classifiers based on AU-ROC (rank 1 indicating the best performing classifier) from right(best) to left(worst) and denotes a lack of significant difference in AUROC by connecting the similar classifiers with a thick horizontal line. Critical Difference Diagram is computed based on the Friedman test and a post-hoc analysis based on the Wilcoxon-Holm method. All P-values are two-sided and significant at P*<*0.05.

### CPT Complexity and Procedure Prevalence Assessment

In order to describe the models’ performance variance for different CPTs, we introduced the CPT complexity measure. The proposed complexity measure is composed of two parts described below. Each of the three approaches (TF-IDF, Doc2Vec, BERT), generate a feature space in which each clinical note is a point in. Neighbors of a point are points closest to it in that feature space based on their Euclidean distance.

*Average same label neighbor ratio*: For each positive note, *M* nearest neighbors were identified, *M* being 10% of overall positive samples for each CPT. (e.g., for CPT 20680 with 4470 positive cases, 447 nearest neighbors were considered and for CPT 25390 with 127 cases, 13 nearest neighbors were considered). The ratio of the same label neighbors within the 10% neighbors was then calculated 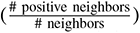. This ratio was calculated for each positive note and then averaged.*Average same label neighbor distance*: For each positive note a sphere was established (with that particular note as the center) to contain 10% of the nearest same label neighbors. The radius of the sphere was then normalized by the longest distance between two positive notes in the dataset. The normalized radius was calculated for every positive note and then averaged.

The two described measures were calculated for each CPT. A higher *average same label neighbor ratio* and a lower *average same label neighbor distance* indicate that the positive samples are better clustered together in the space (more condensed and separated clusters), which in turn makes it easier to identify them from negative ones, meaning it is easier to classify the notes with that CPT. To get a single measure for computing complexity we combined the two measures 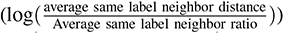.

This measure was used to categorize CPTs (i.e., low complexity, medium complexity, and high complexity), to better investigate the effect of dataset size on model’s performance.

## Results

### Model Comparison/ Performance Evaluation

#### TF-IDF

The AUROC for settings 1, 2, and 3, described previously, were not statistically significant (P*>*0.05), and setting 4 (classified with the K most relevant keywords) showed the best performance, with higher AUROC compared to all other 3 settings (P*<*3.5e-08) (Figure 1). The complete set of results of the best performing setting (4: TF-IDF Feature Selection (Variable K)) in predicting the top 100 CPT codes are presented in Supplementary Tables S1. Figure 4 shows the relevant keywords identified for the 12 most common CPTs based on F-values of the ANOVA test, with larger font representing a greater weight of their F-values (relevance). The presence of the relevant keywords in each word cloud indicates the proper feature selection for each classification task.

**Fig. 1:**
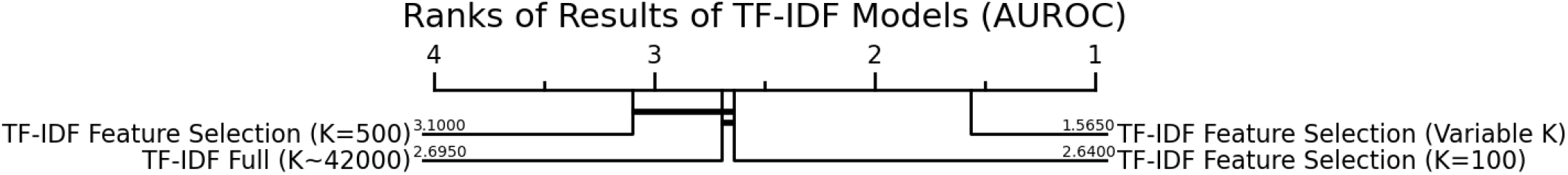
Model performance metrics in different TF-IDF based classifications to predict the top 100 CPT codes. The horizontal connecting lines in the AUROC ranking graphs indicate lack of significant difference between connected groups.

#### Doc2vec

The comparisons in AUROC between different doc2vec settings are shown in Figure 2. As shown in the Critical Difference Diagram, changing the doc2vec parameters did not result in significant changes in model AUROC (the top 3 Doc2vec settings P*>*0.9). The detailed performance metrics for the doc2vec model with the top rank (D=100, W=5, DM) is reported in the Supplementary Table S1.

**Fig. 2:**
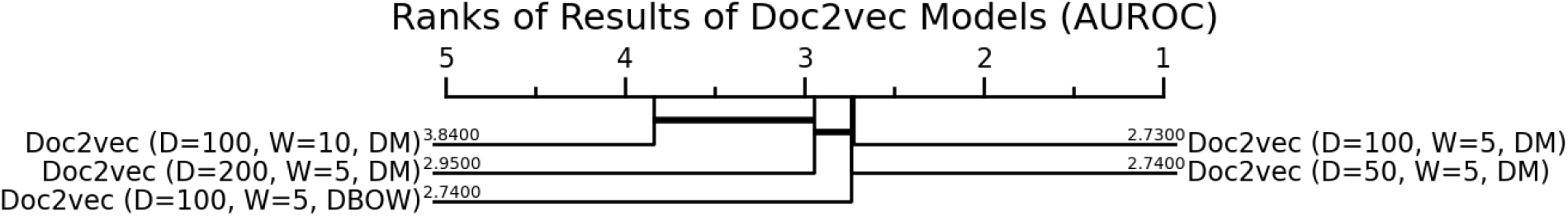
Model performance metrics in different Doc2vec based classifications to predict the top 100 CPT codes. The horizontal connecting lines in the AUROC ranking graphs indicate lack of significant difference between connected groups.

#### BERT

A comparison of the AUROC values for the 5 tested BERT configurations is presented in Figure 3. As shown in the Critical Difference Diagram, there were no differences in AUROC between different BERT configurations (the top 3 BERT settings P*>*0.6). The full results on 100 CPTs for the top rank (A W) are reported in the Supplementary Table S1.

**Fig. 3:**
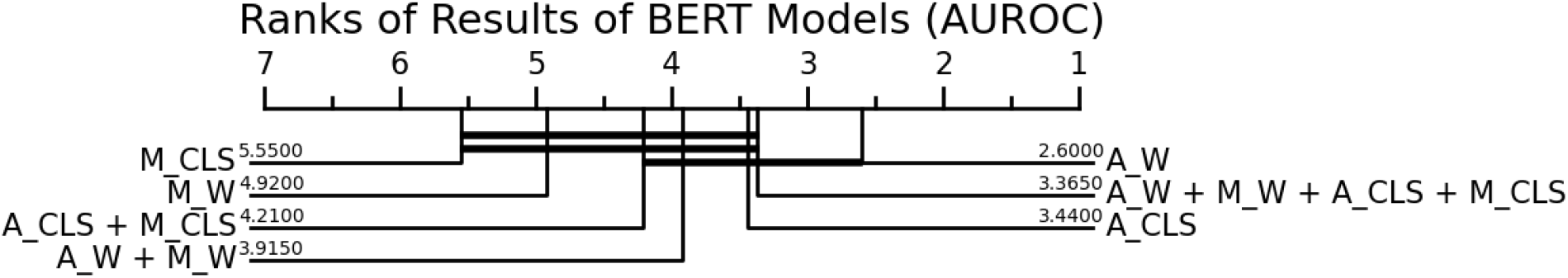
Model performance metrics in different BERT-based classifications to predict the top 100 CPT codes. The horizontal connecting lines in the AUROC ranking graphs indicate lack of significant difference between connected groups.

### Relative Model Performance

The statistical ranking of the AUROC values for the best-performing models for each approach (i.e., TF-IDF, Doc2vec, and BERT) is presented in Figure 5. The TF-IDF approach with varying feature selection had a significantly higher AU-ROC compared to the Doc2vec (by 0.24 ±0.10; P= 1.1e-17) and BERT (by 0.11 ±0.05; P=4.4e-17) approaches (Figure 5). Similar trends were observed in accuracy (P*<*7.0e-08), Sensitivity (P*<*1.04e-15), and Specificity (P*<*5.6e-06), as seen in Figure 6.

**Fig. 4:**
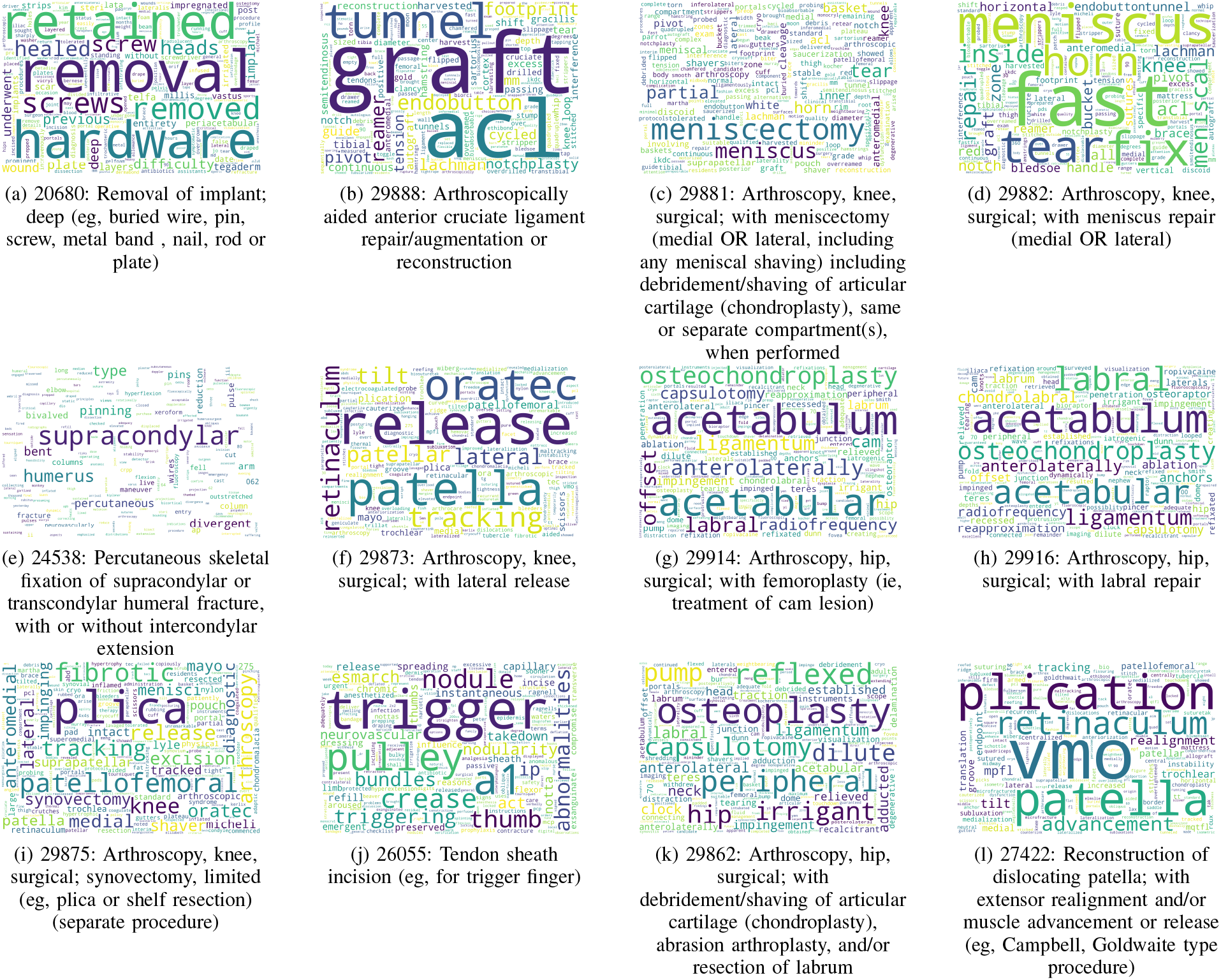
Most important keywords in identifying top 12 most common CPTs and the description of the procedure

**Fig. 5:**
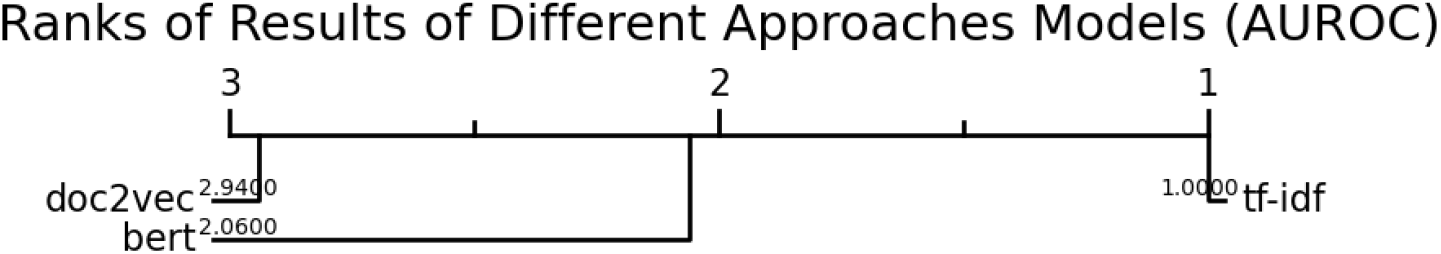
Differences in model performance metrics between best performing TF-IDF, Doc2vec and BERT models to predict the top 100 CPT codes.

**Fig. 6:**
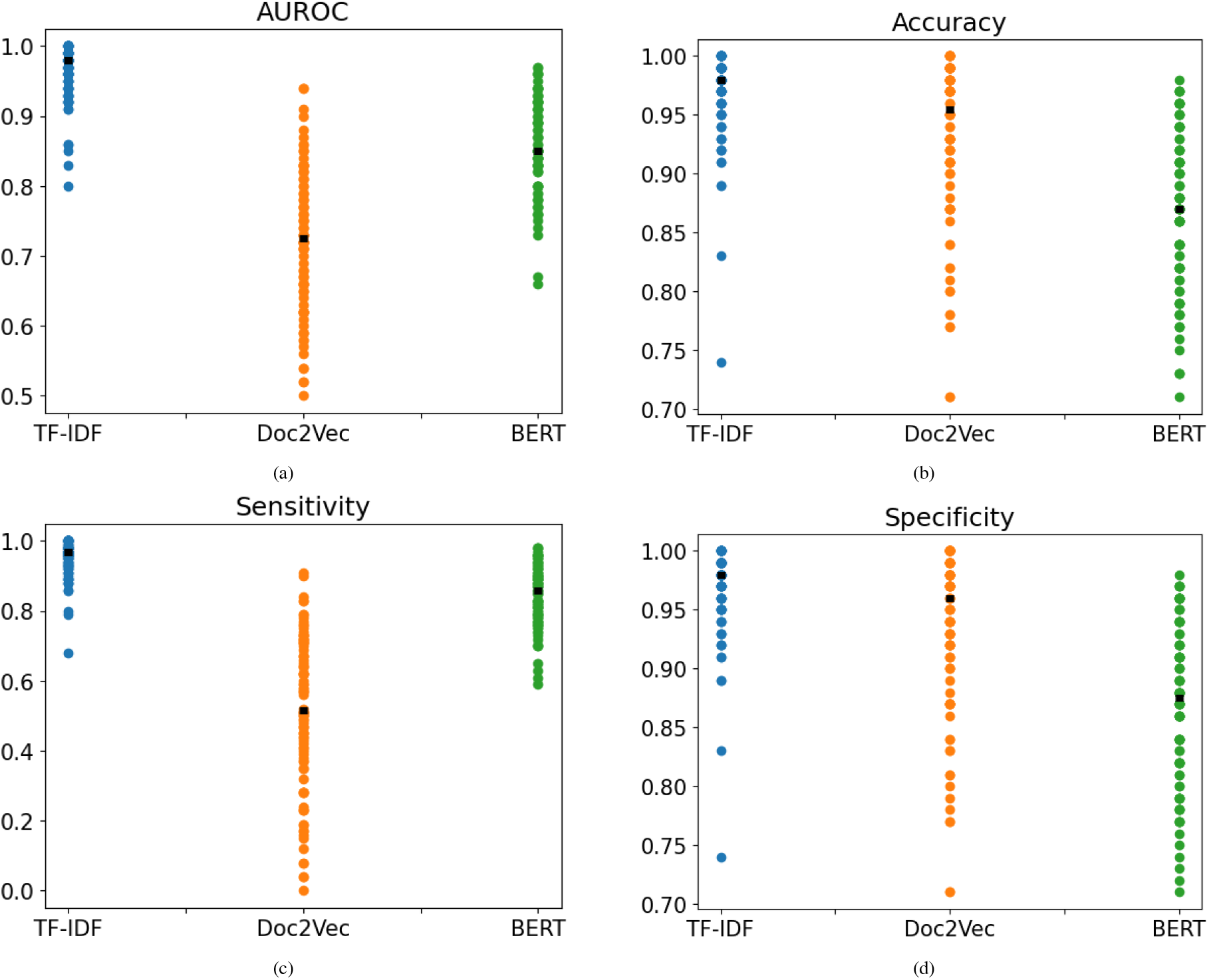
Comparative performance metrics across the three feature extraction models (TF-IDF, Doc2Vec, and BERT). The black dot represents the median.

### Procedure prevalence and CPT Complexity

Considering the superior performance of the TF-IDF approach with variable feature size compared to other approaches, we only investigated the relationships between the CPT complexity and the model performance (i.e., AUROC) for the TF-IDF model. The calculated complexity metrics for each CPT are presented in the Supplementary Table S1. Figure 8 displays the *average ratio* and *distance* measures for CPT 298888 (low complexity) and CPT 29999 (high complexity). Each dot represents an operative note in TF-IDF space which has been projected to 2-dimensional space using principal component analysis (PCA) [33] and TSNE [34] for visualization. CPT 29888 (Arthroscopically Aided Anterior Cruciate Ligament Repair/Augmentation or Reconstruction) is a specific and well-defined surgery; therefore, its corresponding notes are clustered nicely in Figure 8. In contrast, CPT 29999 (Unlisted Procedure, Arthroscopy)is an undefined arthroscopic procedure, which is vague and thus heterogeneous, as observed in the dispersed blue cluster. In Figure 8, the solid purple circle represents the space containing 10% nearest neighbors, used to calculate the average same label ratio. The dotted circles represent the the largest distance in the same label cluster. For CPT29888, the same label ratio is 1, since all the neighbors in its solid purple circle have the same label. Hence, circles representing same label neighbor ratio and distance fall on top of each other. For CPT 29999, the same label ratio is 0; none of the neighbors in the solid purple circle have the same label. For the same label neighbor distance, a larger sphere is required to encompass 10% of the blue samples, which is represented by the dotted black circle. A higher complexity score means the classification problem is more difficult. For example, the complexity scores for CPT codes 29888 and 29999 is -0.34 and 2.33, respectively. Table II shows 5 least and most complex CPTs and their descriptions, and Figure 8 shows the histogram of complexity scores for different CPT codes. As shown in Table II and Supplementary Table S1, most of the CPTs with high complexity do not have a precise definition and are ambiguous. The complexity score is able to describe the variations between AUROC for different CPTs with with adjusted R-squared of 0.611 (P *<* 0.001).

**Fig. 7:**
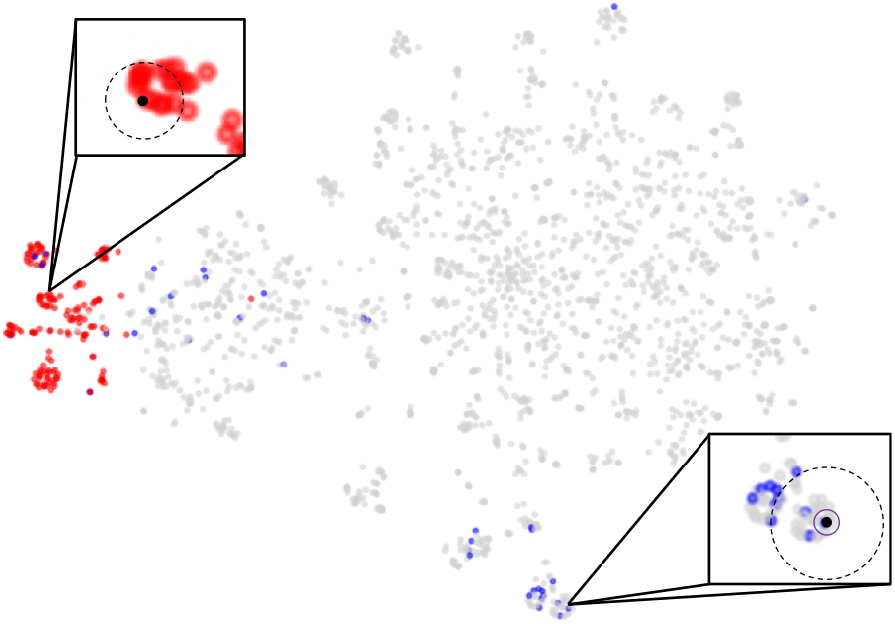
Average ratio and radius measures for complexity for CPT298888 (arthroscopically aided anterior cruciate ligament repair/augmentation or reconstruction; low complexity), seen in red, and CPT29999 (arthroscopy procedures in the musculoskeletal system that do not have a specific code; high complexity), seen in blue. The light gray notes are any procedure other than 29888 and 29999. For cleaner presentation, only a 5% subset of all operative notes is shown.

**Fig. 8:**
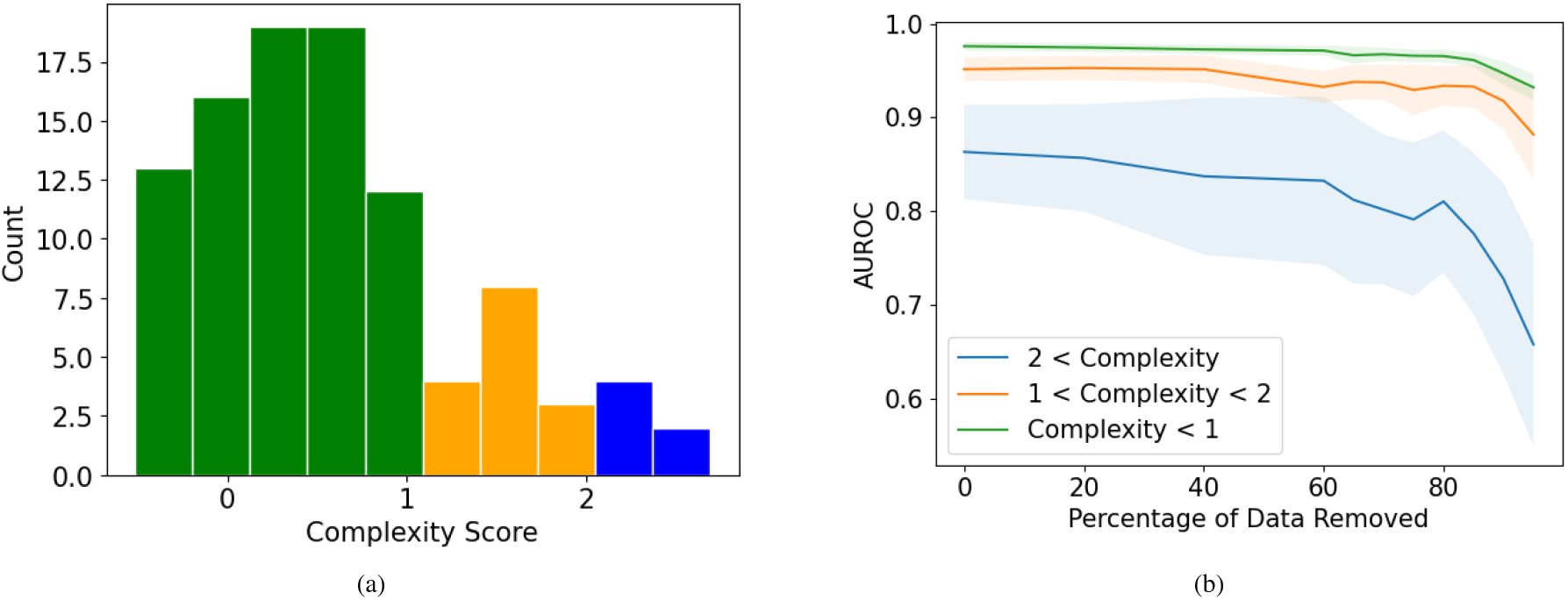
CPT complexity assessment:(a) Histogram of Complexity Scores for our 100 Most Prevalent CPT Codes. (b) Effect of the number of complex notes used during training.

**TABLE I:** Evaluation metrics of the top 12 common CPT codes across the classification models with TF_IDF consistently performing better.

| *CPT* | *BERT* |  |  |  | *Doc2Vec* |  |  |  | *TF_IDF* |  |  |  |
| --- | --- | --- | --- | --- | --- | --- | --- | --- | --- | --- | --- | --- |
|  | Acc | ROC | Spec | Sen | Acc | ROC | Spec | Sen | Acc | ROC | Spec | Sen |
| 20680 | 0.77 | 0.77 | 0.77 | 0.77 | 0.82 | 0.77 | 0.83 | 0.72 | **0.97** | **0.96** | **0.97** | **0.96** |
| 29888 | 0.9 | 0.93 | 0.89 | 0.96 | 0.98 | 0.94 | 0.98 | 0.9 | **1.0** | **0.99** | **1.0** | **0.99** |
| 29881 | 0.78 | 0.83 | 0.77 | 0.88 | 0.91 | 0.72 | **0.93** | 0.51 | **0.92** | **0.95** | 0.92 | **0.98** |
| 29882 | 0.79 | 0.85 | 0.78 | 0.92 | 0.92 | 0.73 | 0.94 | 0.51 | **0.98** | **0.97** | **0.98** | **0.96** |
| 24538 | 0.97 | 0.96 | 0.97 | 0.95 | 0.93 | 0.86 | 0.94 | 0.79 | **0.99** | **0.99** | **0.99** | **0.99** |
| 29873 | 0.89 | 0.88 | 0.89 | 0.86 | 0.95 | 0.79 | 0.96 | 0.62 | **0.98** | **0.97** | **0.98** | **0.97** |
| 29914 | 0.95 | 0.95 | 0.95 | 0.94 | 0.97 | 0.88 | **0.98** | 0.78 | **0.98** | **0.99** | **0.98** | **1.0** |
| 29916 | 0.94 | 0.94 | 0.94 | 0.94 | **0.98** | 0.87 | **0.98** | 0.76 | **0.97** | **0.98** | 0.97 | **1.0** |
| 29875 | 0.8 | 0.8 | 0.8 | 0.79 | 0.81 | 0.78 | 0.81 | 0.75 | **0.94** | **0.96** | **0.94** | **0.98** |
| 26055 | 0.94 | 0.9 | 0.94 | 0.86 | **0.98** | 0.84 | **0.98** | 0.7 | **1.0** | **0.98** | **1.0** | **0.97** |
| 29862 | 0.92 | 0.94 | 0.92 | 0.96 | **0.97** | 0.82 | **0.98** | 0.65 | 0.96 | **0.98** | 0.96 | **1.0** |
| 27422 | 0.91 | 0.91 | 0.91 | 0.91 | **0.98** | 0.75 | **0.99** | 0.5 | **0.98** | **0.98** | **0.98** | **0.99** |

**TABLE II:** CPT Complexity: The tables lists the top 5 least and most common CPTs along with their descriptions.

| *CPT* | *Complexity* | *CPT Description* |
| --- | --- | --- |
| 20615 | -0.51 | Aspiration and injection for treatment of bone cyst |
| 26055 | -0.47 | Tendon sheath incision (eg, for trigger finger) |
| 29914 | -0.42 | Arthroscopy, hip, surgical; with femoroplasty (ie, treatment of cam lesion) |
| 25111 | -0.36 | Excision of ganglion, wrist (dorsal or volar); primary |
| 21320 | -0.35 | Closed treatment of nasal bone fracture; with stabilization |
| 20900 | 2.16 | Bone graft, any donor area; minor or small (eg, dowel or button) |
| 29065 | 2.28 | Application, cast; shoulder to hand (long arm) |
| 29999 | 2.33 | Unlisted procedure, arthroscopy |
| 27599 | 2.45 | Unlisted procedure, femur or knee |
| 27899 | 2.69 | Unlisted procedure, leg or ankle |

In this study, we look at 100 most common CPTs with their frequency ranging from 127 (CPT 25390) to 4470 (CPT 20680). The classification performance for predicting the CPT codes using the notes were on average relatively high but had a wide range (average AUROC: 0.96 ±0.04, average accuracy: 0.97 ±0.04, average specificity: 0.97 ±0.04, average sensitivity: 0.96 ±0.05 these results are based on TF-IDF with feature selection, the best performing model). Considering the high degrees of variability in CPT frequencies, a question arises is that how do these results change by changing the amount of training data and whether this relationship is affected by CPT complexity. To assess the effect of procedure prevalence (count) on the model’s ability to predict the CPT, we randomly removed the notes for each procedure (CPT) by up to 90% with 10% increments and then used the reduced data to train the model and predict the CPT codes. This analysis was only done for the best-performing prediction model. The CPT codes were grouped to low complexity (complexity score*<*2), medium complexity (1 ⩽ Complexity Score ⩽ 2) and complex (Complexity score*>*2). The changes in AUROC for best performing approach (TF-IDF with feature selection) relative to random reduction in data are presented (Figure 8). For low to medium complexity CPTs, the AUROC dropped only after extreme (*>*80%) reductions in the number of notes. However, for complex CPTs, the was a continuous decline in AUROC by reductions in number of notes with sharpest declines after 80% reduction.

### Quality Assessment

In addition to being used for automating CPT code assignment, the approaches in this study can be used for quality assessment in the CPT assignment process. As an example, we looked at CPT 29888 (arthroscopically aided anterior cruciate ligament repair/augmentation or reconstruction), which is one of the most common orthopedic procedures and the second most frequent CPT code in our dataset. The TF-IDF model was able to predict CPT 29888 with AUROC of 0.99, accuracy of 1, specificity of 1 and sensitivity of 0.99. Following 5-fold cross validation, there were 203 incorrect predictions out of 44,002 operative notes. An independent examiner reviewed and relabeled the mislabeled notes, blinded to the ground truth or the predicted label. Of the 203 mislabeled notes, 198 (97.5%) notes were classified correctly by the model and had incorrect ground truth labels. Three note were incomplete, and 2 notes were truly misclassified by model. Hence, the model can be used for quality assessment and to identify incorrect CPT assignments.

## Discussion

In this study, we systematically analyzed the ability of three commonly used NLP models to predict the 100 most common musculoskeletal CPT codes from unstructured operative notes. Our findings rejects our first hypothesis as the traditional TF-IDF model with a dynamic feature space size outperformed the more fine-tuned, computationally expensive BERT model. Our results support our second hypothesis demonstrating that CPT complexity can explain up to 61% variability in AUROC. Finally, we saw that prevalence of procedures (CPT counts) only influence the model’s prediction performance in complex cases.

This study showed that NLP models can perform with near-human accuracy, and even highlight human error. The AUROC of the top-performing TF-IDF model was 0.99 for arthroscopic ACL surgery (CPT 29888), a common procedure with relatively low CPT complexity. In that procedure alone, 97.5% of the codes thought to be misclassified by the model were found to have incorrect ground truth codes. While we did not assess the misclassification of the entire corpus, other studies, such as one automating the classification of anesthesia CPT coding by Burns et al [5] have highlighted a manual CPT assignment error rate of 5.0%, a significant proportion that undoubtedly affects insurance claims and hospital revenue. The performance of our models is similar to that of Levy et al [10], who characterized over 93,000 pathology report CPT codes using SVM, BERT, and Extreme Gradient Boosting Trees (XGBoost). Their group demonstrated that XGBoost and BERT models yielded comparable, promising results with XGBoost slightly outperforming BERT. Similarly, Joo et al [35] used a two-step neural machine translation (NMT) model, which they compared to SVM and LSTM models, to automate the classification of anesthesia procedure CPT codes using over 187,000 documented procedures. They observed promising and similar performance in all models. The implications of these studies, as well as ours, is that there is significant utility in using these NLP models for billing processes in healthcare systems, including quality control of insurance claims. Currently, the financial impact of billing and insurance related tasks is estimated to be around $25 billion USD [2], [36]–[38], and the Centers for Medicare and Medicaid Services reported that improper payments amounted to $28.91 billion USD in 2019 alone [39]. The concept of automating billing processes to reduce spending and staffing is not new; in 2006, Pakhomov et al [40] documented their development and implementation of an automated ICD coding system that was able to code 48% of problem list entries with 98% precision. This system resulted in the reduction of staff from 34 coders to 7 verifiers. Now, as we have proof of concept that these models work with CPT codes, and that they have been successfully implemented in the past for other coding processes, it is important in that we continue to augment our currently flawed systems with automated and/or assisted CPT prediction systems.

Interestingly, in this study our best performing TF-IDF model outperformed our best performing BERT model, with both models outperforming the Doc2vec models. This is similar to the previous findings. Kim et al [2], who sought to predict CPT codes using spinal surgery operative notes with a long short-term memory (LSTM) deep learning model and a random forest model. In their analysis 391 operative notes and 15 unique CPT codes, their random forest model outperformed their LSTM deep learning model with AUROCs of 0.94 and 0.72, respectively. Their group was also expecting the more advanced LSTM model to outperform their traditional random forest model. They proposed that with larger sample size, they would expect deep learning to outperform random forest model. Levy et al [10] found that their XGBoost outperformed BERT models in CPT classification. Since bag-of-word features were fed to XGBoost, and XGBoost basically has feature selection within itself, it is more similar to our best performing model, TF-IDF with dynamic feature space. Few studies report the efficacy of Doc2vec in coding automation processes [3].Li et al [9] proposed an ICD-9 coding method using a deep learning framework called DeepLabeler, which combines a convoluted neural network (CNN) with Doc2vec to assign ICD codes. They concluded that Doc2vec was critical to their prediction accuracy, and their DeepLabeler outperformed both hierarchy-based and flat-SVM approaches.

The superiority of traditional models to deep learning models in these tasks could be attributed to few factors. Deep learning models generally need much more data to train compared to traditional approaches, and in clinical domain, finding bigger datasets is very difficult because of privacy and security concerns and manual labeling which can also be very expensive. Another challenge in working with clinical notes is the presence of noise and also templates. Physicians who write these notes tend to keep a template for their surgeries and only change details in the template each time. Generally each physician has their own template which considerably affects the note representations however does not give much information as to the procedure itself, but since traditional approaches take keywords into account they are less impacted by these templates. The same is for the noise in the data. Clinical notes are very noisy, they are usually written in a rush, hence they have a lot of misspellings, incorrect grammar, acronyms, etc. Also in many cases these notes are actually forms that are saved in text format, so there are a lot of white space and non-ASCII characters included in the text. While there are different variations of BERT models, pre-trained on different datasets, all of them are clean formatted datasets which are very different from clinical notes.

Another outcome of this study is that the number of notes available for a well-defined CPT in the data set, do not significantly affect the results. This information alleviates the burden of collecting large data sets for simple cohort identifications. If there are enough samples from a cohort, gathering more data is not going to improve the results significantly. It is worth noting that the least frequent CPT included in our experiments is 25390 CPT with frequency of 127 has a AUROC value of 0.99 with TF-IDF model, hence the threshold for having enough number of samples can be quite low.

This study is not without limitations. In this study, we highlighted CPT 29888 as an example of human error in the code assignment process. As our training sets were derived from data with manual CPT errors, we expect that models trained on these sets would have their own inherent flaws. Additionally, the operative reports in this study were all from the same institution. As such, it is possible that bias from overfitting may exist due to billing practices unique to our institution.

## Conclusion

The current study supplements the existing literature in support of using NLP to automate codifying clinical notes and to conduct quality control. Results also support use of traditional NLP approaches (i.e., TF-IDF) as proper tools for these well defined applications. The fact that these traditional approaches are less resource intensive compared to the state-of-the-art models such as BERT, lowers the barriers to wide-spread clinical adoption.

## Data Availability

The data used in this study cannot be publicly shared due to privacy concerns. Data can be access upon reasonable request pending approvals from Boston Children's Hospital.

