## Supplement Table 1 for "Systematic Evaluation of Common Natural Language Processing Techniques to Codify Clinical Notes"

**Table S1.** Model performance metrics for predicting the top 100 common CPT codes from operative notes.

| CPT Code | Procedure Description per CPT Code | Count | CPT Complexity Score | Performance Metrics for TF-IDF | | | | | Performance Metrics for Doc2Vec | | | | Performance Metrics for BERT | | | |
|---|---|---|---|---|---|---|---|---|---|---|---|---|---|---|---|---|
| | | | | AUROC | Accuracy | Specificity | Sensetivity | Number of Features | AUROC | Accuracy | Specificity | Sensetivity | AUROC | Accuracy | Specificity | Sensetivity |
| 20680 | Removal of implant; deep (eg, buried wire, pin, screw, metal band, nail, rod or plate) | 4470 | 0.49 | 0.96 | 0.97 | 0.97 | 0.96 | 630 | 0.77 | 0.82 | 0.83 | 0.72 | 0.77 | 0.77 | 0.77 | 0.77 |
| 29888 | Arthroscopically aided anterior cruciate ligament repair/augmentation or reconstruction | 3727 | -0.34 | 0.99 | 1 | 1 | 0.99 | 630 | 0.94 | 0.98 | 0.98 | 0.9 | 0.93 | 0.9 | 0.89 | 0.96 |
| 29881 | Arthroscopy, knee, surgical; with meniscectomy (medial OR lateral, including any meniscal shaving) including debridement/shaving of articular cartilage (chondroplasty), same or separate compartment(s), when performed | 2032 | 0.54 | 0.95 | 0.92 | 0.92 | 0.98 | 460 | 0.72 | 0.91 | 0.93 | 0.51 | 0.83 | 0.78 | 0.77 | 0.88 |
| 29882 | Arthroscopy, knee, surgical; with meniscus repair (medial OR lateral) | 1985 | 0.49 | 0.97 | 0.98 | 0.98 | 0.96 | 630 | 0.73 | 0.92 | 0.94 | 0.51 | 0.85 | 0.79 | 0.78 | 0.92 |
| 24538 | Percutaneous skeletal fixation of supracondylar or transcondylar humeral fracture, with or without intercondylar extension | 1656 | 0 | 0.99 | 0.99 | 0.99 | 0.99 | 460 | 0.86 | 0.93 | 0.94 | 0.79 | 0.96 | 0.97 | 0.97 | 0.95 |
| 29873 | Arthroscopy, knee, surgical; with lateral release | 1492 | 0.17 | 0.97 | 0.98 | 0.98 | 0.97 | 460 | 0.79 | 0.95 | 0.96 | 0.62 | 0.88 | 0.89 | 0.89 | 0.86 |
| 29914 | Arthroscopy, hip, surgical; with femoroplasty (ie, treatment of cam lesion) | 1435 | -0.42 | 0.99 | 0.98 | 0.98 | 1 | 910 | 0.88 | 0.97 | 0.98 | 0.78 | 0.95 | 0.95 | 0.95 | 0.94 |
| 29916 | Arthroscopy, hip, surgical; with labral repair | 1065 | -0.33 | 0.98 | 0.97 | 0.97 | 1 | 460 | 0.87 | 0.98 | 0.98 | 0.76 | 0.94 | 0.94 | 0.94 | 0.94 |
| 29875 | Arthroscopy, knee, surgical; synovectomy, limited (eg, plica or shelf resection) (separate procedure) | 1021 | 0.77 | 0.96 | 0.94 | 0.94 | 0.98 | 910 | 0.78 | 0.81 | 0.81 | 0.75 | 0.8 | 0.8 | 0.8 | 0.79 |
| 26055 | Tendon sheath incision (eg, for trigger finger) | 868 | -0.47 | 0.98 | 1 | 1 | 0.97 | 460 | 0.84 | 0.98 | 0.98 | 0.7 | 0.9 | 0.94 | 0.94 | 0.86 |
| 29862 | Arthroscopy, hip, surgical; with debridement/shaving of articular cartilage (chondroplasty), abrasion arthroplasty, and/or resection of labrum | 820 | -0.06 | 0.98 | 0.96 | 0.96 | 1 | 510 | 0.82 | 0.97 | 0.98 | 0.65 | 0.94 | 0.92 | 0.92 | 0.96 |
| 27422 | Reconstruction of dislocating patella; with extensor realignment and/or muscle advancement or release (eg, Campbell, Goldwaite type procedure) | 802 | 0.14 | 0.98 | 0.98 | 0.98 | 0.99 | 460 | 0.75 | 0.98 | 0.99 | 0.5 | 0.91 | 0.91 | 0.91 | 0.91 |
| 29999 | Unlisted procedure, arthroscopy | 713 | 2.33 | 0.83 | 0.74 | 0.74 | 0.92 | 460 | 0.7 | 0.78 | 0.79 | 0.62 | 0.77 | 0.73 | 0.72 | 0.81 |
| 20670 | Removal of implant; superficial (eg, buried wire, pin or rod) (separate procedure) | 658 | 1.75 | 0.92 | 0.91 | 0.91 | 0.93 | 80 | 0.77 | 0.77 | 0.77 | 0.77 | 0.78 | 0.79 | 0.79 | 0.77 |
| 29879 | Arthroscopy, knee, surgical; abrasion arthroplasty (includes chondroplasty where necessary) or multiple drilling or microfracture | 653 | 0.92 | 0.95 | 0.94 | 0.94 | 0.96 | 910 | 0.77 | 0.86 | 0.86 | 0.67 | 0.83 | 0.84 | 0.84 | 0.82 |
| 22899 | Unlisted procedure, spine | 629 | 0.08 | 0.99 | 0.99 | 0.99 | 0.98 | 910 | 0.85 | 0.97 | 0.98 | 0.73 | 0.92 | 0.94 | 0.94 | 0.89 |
| 29806 | Arthroscopy, shoulder, surgical; capsulorrhaphy | 620 | -0.33 | 1 | 0.99 | 0.99 | 1 | 910 | 0.83 | 0.99 | 1 | 0.66 | 0.96 | 0.97 | 0.97 | 0.96 |
| 29877 | Arthroscopy, knee, surgical; debridement/shaving of articular cartilage (chondroplasty) | 521 | 1.52 | 0.94 | 0.92 | 0.92 | 0.96 | 910 | 0.78 | 0.87 | 0.87 | 0.69 | 0.83 | 0.84 | 0.84 | 0.82 |
| 29886 | Arthroscopy, knee, surgical; drilling for intact osteochondritis dissecans lesion | 487 | 0.53 | 0.97 | 0.97 | 0.97 | 0.97 | 910 | 0.81 | 0.89 | 0.89 | 0.72 | 0.86 | 0.84 | 0.84 | 0.89 |
| 26727 | Percutaneous skeletal fixation of unstable phalangeal shaft fracture, proximal or middle phalanx, finger or thumb, with manipulation, each | 436 | -0.04 | 0.99 | 0.99 | 0.99 | 1 | 460 | 0.71 | 0.98 | 0.99 | 0.43 | 0.93 | 0.91 | 0.91 | 0.95 |
| 29915 | Arthroscopy, hip, surgical; with acetabuloplasty (ie, treatment of pincer lesion) | 406 | 0.41 | 0.98 | 0.96 | 0.95 | 1 | 460 | 0.73 | 0.98 | 0.99 | 0.47 | 0.94 | 0.94 | 0.94 | 0.94 |
| 27685 | Lengthening or shortening of tendon, leg or ankle; single tendon (separate procedure) | 397 | 0.87 | 0.96 | 0.97 | 0.97 | 0.95 | 910 | 0.76 | 0.9 | 0.9 | 0.62 | 0.85 | 0.82 | 0.82 | 0.89 |
| 29887 | Arthroscopy, knee, surgical; drilling for intact osteochondritis dissecans lesion with internal fixation | 397 | 0.33 | 0.98 | 0.98 | 0.98 | 0.97 | 910 | 0.82 | 0.91 | 0.91 | 0.73 | 0.84 | 0.86 | 0.87 | 0.81 |
| 27095 | Injection procedure for hip arthrography; with anesthesia | 375 | 0.25 | 0.98 | 0.99 | 0.99 | 0.96 | 910 | 0.83 | 0.92 | 0.92 | 0.73 | 0.83 | 0.88 | 0.88 | 0.79 |
| 27475 | Arrest, epiphyseal, any method (eg, epiphysiodesis); distal femur | 373 | 0.54 | 0.99 | 0.98 | 0.98 | 1 | 910 | 0.64 | 0.99 | 1 | 0.28 | 0.85 | 0.84 | 0.84 | 0.85 |
| 27420 | Reconstruction of dislocating patella; (eg, Hauser type procedure) | 363 | 1.07 | 0.98 | 0.96 | 0.96 | 1 | 910 | 0.76 | 0.93 | 0.94 | 0.58 | 0.91 | 0.9 | 0.89 | 0.93 |
| 24575 | Open treatment of humeral epicondylar fracture, medial or lateral, includes internal fixation, when performed | 354 | 0.06 | 0.98 | 0.99 | 0.99 | 0.97 | 910 | 0.69 | 0.87 | 0.87 | 0.51 | 0.84 | 0.8 | 0.8 | 0.87 |
| 29874 | Arthroscopy, knee, surgical; for removal of loose body or foreign body (eg, osteochondritis dissecans fragmentation, chondral fragmentation) | 342 | 1.5 | 0.92 | 0.96 | 0.96 | 0.88 | 460 | 0.71 | 0.87 | 0.87 | 0.56 | 0.82 | 0.83 | 0.83 | 0.81 |
| 27418 | Anterior tibial tubercleplasty (eg, Maquet type procedure) | 333 | 0.23 | 1 | 0.99 | 0.99 | 1 | 460 | 0.83 | 0.95 | 0.95 | 0.72 | 0.89 | 0.91 | 0.91 | 0.87 |
| 29863 | Arthroscopy, hip, surgical; with synovectomy | 324 | 0.44 | 0.98 | 0.97 | 0.97 | 1 | 910 | 0.68 | 0.99 | 1 | 0.37 | 0.94 | 0.92 | 0.92 | 0.95 |
| 28116 | Ostectomy, excision of tarsal coalition | 317 | -0.23 | 0.99 | 1 | 1 | 0.98 | 130 | 0.58 | 0.99 | 1 | 0.16 | 0.9 | 0.86 | 0.86 | 0.94 |
| 25606 | Percutaneous skeletal fixation of distal radial fracture or epiphyseal separation | 315 | 0.6 | 0.98 | 0.98 | 0.98 | 0.97 | 910 | 0.78 | 0.84 | 0.84 | 0.71 | 0.82 | 0.79 | 0.79 | 0.86 |
| 29895 | Arthroscopy, ankle (tibiotalar and fibulotalar joints), surgical; synovectomy, partial | 314 | 0.16 | 0.98 | 0.98 | 0.98 | 0.98 | 460 | 0.75 | 0.93 | 0.93 | 0.57 | 0.91 | 0.91 | 0.91 | 0.9 |
| 29884 | Arthroscopy, knee, surgical; with lysis of adhesions, with or without manipulation (separate procedure) | 313 | 0.47 | 0.97 | 0.99 | 0.99 | 0.95 | 80 | 0.74 | 0.91 | 0.91 | 0.57 | 0.8 | 0.84 | 0.84 | 0.76 |
| 25111 | Excision of ganglion, wrist (dorsal or volar); primary | 309 | -0.36 | 0.99 | 1 | 1 | 0.98 | 80 | 0.62 | 0.99 | 0.99 | 0.24 | 0.87 | 0.88 | 0.88 | 0.85 |
| 27635 | Excision or curettage of bone cyst or benign tumor, tibia or fibula; | 300 | 0.48 | 0.94 | 0.97 | 0.97 | 0.92 | 510 | 0.63 | 0.87 | 0.87 | 0.38 | 0.8 | 0.87 | 0.87 | 0.72 |
| 25575 | Open treatment of radial AND ulnar shaft fractures, with internal fixation, when performed; of radius AND ulna | 286 | 0.46 | 0.99 | 0.99 | 0.99 | 1 | 460 | 0.71 | 0.93 | 0.93 | 0.49 | 0.85 | 0.82 | 0.82 | 0.88 |
| 27606 | Tenotomy, percutaneous, Achilles tendon (separate procedure); general anesthesia | 285 | 0.88 | 0.97 | 0.98 | 0.98 | 0.96 | 80 | 0.65 | 0.99 | 0.99 | 0.32 | 0.84 | 0.87 | 0.87 | 0.81 |
| 20615 | Aspiration and injection for treatment of bone cyst | 267 | -0.51 | 0.99 | 1 | 1 | 0.98 | 910 | 0.8 | 1 | 1 | 0.6 | 0.89 | 0.96 | 0.96 | 0.83 |
| 29861 | Arthroscopy, hip, surgical; with removal of loose body or foreign body | 266 | -0.2 | 0.97 | 0.96 | 0.96 | 0.98 | 910 | 0.94 | 0.97 | 0.97 | 0.91 | 0.96 | 0.96 | 0.96 | 0.96 |
| 27485 | Arrest, hemiepiphyseal, distal femur or proximal tibia or fibula (eg, genu varus or valgus) | 266 | 0.65 | 0.96 | 0.98 | 0.98 | 0.94 | 80 | 0.57 | 0.99 | 1 | 0.15 | 0.9 | 0.84 | 0.84 | 0.96 |
| 20610 | Arthrocentesis, aspiration and/or injection; major joint or bursa (eg, shoulder, hip, knee joint, subacromial bursa) | 263 | 1.05 | 0.94 | 0.97 | 0.97 | 0.91 | 460 | 0.79 | 0.95 | 0.95 | 0.64 | 0.82 | 0.88 | 0.88 | 0.75 |
| 29010 | Application of Risser jacket, localizer, body; only | 263 | -0.07 | 1 | 1 | 1 | 1 | 460 | 0.81 | 0.99 | 0.99 | 0.62 | 0.97 | 0.96 | 0.96 | 0.98 |
| 20694 | Removal, under anesthesia, of external fixation system | 255 | 0.21 | 1 | 1 | 1 | 1 | 460 | 0.77 | 0.82 | 0.83 | 0.71 | 0.85 | 0.89 | 0.89 | 0.82 |
| 29807 | Arthroscopy, shoulder, surgical; repair of SLAP lesion | 247 | 0.21 | 0.99 | 0.98 | 0.98 | 1 | 130 | 0.85 | 0.97 | 0.97 | 0.73 | 0.97 | 0.97 | 0.97 | 0.98 |
| 29846 | Arthroscopy, wrist, surgical; excision and/or repair of triangular fibrocartilage and/or joint debridement | 244 | -0.08 | 0.99 | 1 | 1 | 0.98 | 910 | 0.68 | 1 | 1 | 0.37 | 0.85 | 0.81 | 0.81 | 0.9 |
| 27605 | Tenotomy, percutaneous, Achilles tendon (separate procedure); local anesthesia | 243 | -0.06 | 1 | 0.99 | 0.99 | 1 | 80 | 0.85 | 0.99 | 1 | 0.71 | 0.96 | 0.95 | 0.95 | 0.96 |
| 29405 | Application of short leg cast (below knee to toes); | 238 | 1.43 | 0.92 | 0.89 | 0.89 | 0.96 | 90 | 0.52 | 0.99 | 1 | 0.04 | 0.83 | 0.82 | 0.82 | 0.83 |
| 27355 | Excision or curettage of bone cyst or benign tumor of femur; | 235 | 0.26 | 0.93 | 0.95 | 0.95 | 0.91 | 90 | 0.66 | 0.91 | 0.92 | 0.4 | 0.84 | 0.93 | 0.93 | 0.74 |
| 28238 | Reconstruction (advancement), posterior tibial tendon with excision of accessory tarsal navicular bone (eg, Kidner type procedure) | 231 | -0.34 | 0.97 | 1 | 1 | 0.93 | 80 | 0.59 | 0.99 | 1 | 0.17 | 0.92 | 0.95 | 0.95 | 0.89 |
| 23515 | Open treatment of clavicular fracture, includes internal fixation, when performed | 229 | -0.25 | 1 | 1 | 1 | 1 | 910 | 0.66 | 0.93 | 0.94 | 0.39 | 0.94 | 0.97 | 0.97 | 0.91 |
| 27427 | Ligamentous reconstruction (augmentation), knee; extra-articular | 226 | 0.98 | 0.93 | 0.98 | 0.98 | 0.89 | 910 | 0.75 | 0.98 | 0.98 | 0.51 | 0.92 | 0.87 | 0.87 | 0.98 |
| 27827 | Open treatment of fracture of weight bearing articular surface/portion of distal tibia (eg, pilon or tibial plafond), with internal fixation, when performed; of tibia only | 220 | 0.48 | 0.94 | 0.99 | 0.99 | 0.89 | 910 | 0.66 | 0.91 | 0.92 | 0.41 | 0.76 | 0.82 | 0.82 | 0.7 |
| 27687 | Gastrocnemius recession (eg, Strayer procedure) | 217 | 0.62 | 0.97 | 0.98 | 0.98 | 0.95 | 510 | 0.71 | 0.98 | 0.98 | 0.44 | 0.89 | 0.88 | 0.88 | 0.91 |
| 26561 | Repair of syndactyly (web finger) each web space; with skin flaps and grafts | 213 | 0.1 | 0.99 | 0.99 | 0.99 | 0.98 | 910 | 0.62 | 0.99 | 1 | 0.23 | 0.91 | 0.93 | 0.93 | 0.88 |
| 20552 | Injection(s); single or multiple trigger point(s), 1 or 2 muscle(s) | 212 | 0.23 | 0.98 | 0.99 | 0.99 | 0.98 | 910 | 0.91 | 0.99 | 0.99 | 0.83 | 0.92 | 0.96 | 0.96 | 0.88 |
| 29883 | Arthroscopy, knee, surgical; with meniscus repair (medial AND lateral) | 210 | 1.53 | 0.98 | 0.96 | 0.96 | 1 | 910 | 0.83 | 0.94 | 0.94 | 0.71 | 0.87 | 0.84 | 0.84 | 0.9 |
| 26587 | Reconstruction of polydactylous digit, soft tissue and bone | 204 | 0.04 | 0.98 | 0.99 | 0.99 | 0.98 | 80 | 0.9 | 0.97 | 0.97 | 0.83 | 0.89 | 0.91 | 0.91 | 0.88 |
| 28120 | Partial excision (craterization, saucerization, sequestrectomy, or diaphysectomy) bone (eg, osteomyelitis or bossing); talus or calcaneus | 202 | 0.25 | 0.96 | 0.98 | 0.98 | 0.95 | 910 | 0.54 | 0.99 | 1 | 0.07 | 0.85 | 0.91 | 0.91 | 0.8 |
| 21320 | Closed treatment of nasal bone fracture; with stabilization | 200 | -0.35 | 1 | 1 | 1 | 1 | 940 | 0.72 | 0.99 | 1 | 0.45 | 0.92 | 0.94 | 0.94 | 0.9 |
| 29880 | Arthroscopy, knee, surgical; with meniscectomy (medial AND lateral, including any meniscal shaving) including debridement/shaving of articular cartilage (chondroplasty), same or separate compartment(s), when performed | 196 | 1.6 | 0.95 | 0.93 | 0.93 | 0.97 | 460 | 0.82 | 0.84 | 0.84 | 0.79 | 0.85 | 0.82 | 0.82 | 0.87 |
| 27698 | Repair, secondary, disrupted ligament, ankle, collateral (eg, Watson-Jones procedure) | 193 | 0.25 | 1 | 0.99 | 0.99 | 1 | 510 | 0.76 | 0.87 | 0.87 | 0.64 | 0.84 | 0.89 | 0.89 | 0.79 |

| | | | | | | | | | | | | | | | | |
|---|---|---|---|---|---|---|---|---|---|---|---|---|---|---|---|---|
| 27600 | Decompression fasciotomy, leg; anterior and/or lateral compartments only | 192 | 0.44 | 0.99 | 0.99 | 0.99 | 1 | 510 | 0.72 | 0.96 | 0.96 | 0.47 | 0.83 | 0.9 | 0.9 | 0.76 |
| 24579 | Open treatment of humeral condylar fracture, medial or lateral, includes internal fixation, when performed | 190 | 1.11 | 0.97 | 0.99 | 0.99 | 0.95 | 910 | 0.68 | 0.92 | 0.92 | 0.45 | 0.78 | 0.77 | 0.77 | 0.79 |
| 27096 | Injection procedure for sacroiliac joint, anesthetic/steroid, with image guidance (fluoroscopy or CT) including arthrography when performed | 186 | -0.1 | 0.98 | 0.99 | 0.99 | 0.97 | 510 | 0.81 | 1 | 1 | 0.62 | 0.93 | 0.98 | 0.98 | 0.89 |
| 26735 | Open treatment of phalangeal shaft fracture, proximal or middle phalanx, finger or thumb, includes internal fixation, when performed, each | 186 | 0.77 | 0.92 | 0.98 | 0.98 | 0.86 | 910 | 0.83 | 0.91 | 0.91 | 0.76 | 0.78 | 0.86 | 0.86 | 0.7 |
| 29876 | Arthroscopy, knee, surgical; synovectomy, major, 2 or more compartments (eg, medial or lateral) | 183 | 1.73 | 0.94 | 0.95 | 0.95 | 0.92 | 510 | 0.65 | 0.95 | 0.95 | 0.35 | 0.77 | 0.78 | 0.78 | 0.76 |
| 21235 | Graft; ear cartilage, autogenous, to nose or ear (includes obtaining graft) | 183 | -0.13 | 1 | 1 | 1 | 1 | 910 | 0.59 | 1 | 1 | 0.19 | 0.94 | 0.96 | 0.96 | 0.92 |
| 28344 | Reconstruction, toe(s); polydactyly | 183 | 0.08 | 0.98 | 0.99 | 0.99 | 0.97 | 500 | 0.73 | 0.97 | 0.97 | 0.49 | 0.91 | 0.9 | 0.9 | 0.92 |
| 24545 | Open treatment of humeral supracondylar or transcondylar fracture, includes internal fixation, when performed; without intercondylar extension | 180 | 1.77 | 0.97 | 0.94 | 0.94 | 1 | 500 | 0.72 | 0.78 | 0.78 | 0.67 | 0.88 | 0.92 | 0.92 | 0.83 |
| 29870 | Arthroscopy, knee, diagnostic, with or without synovial biopsy (separate procedure) | 178 | 1.76 | 0.86 | 0.83 | 0.83 | 0.89 | 500 | 0.61 | 0.93 | 0.93 | 0.28 | 0.77 | 0.78 | 0.78 | 0.75 |
| 24999 | Unlisted procedure, humerus or elbow | 177 | 0.99 | 0.91 | 0.96 | 0.96 | 0.86 | 910 | 0.8 | 0.87 | 0.87 | 0.74 | 0.8 | 0.77 | 0.77 | 0.83 |
| 27602 | Decompression fasciotomy, leg; anterior and/or lateral, and posterior compartment(s) | 168 | -0.02 | 0.97 | 0.99 | 0.99 | 0.94 | 300 | 0.56 | 1 | 1 | 0.12 | 0.87 | 0.95 | 0.96 | 0.79 |
| 27599 | Unlisted procedure, femur or knee | 168 | 2.45 | 0.8 | 0.93 | 0.93 | 0.68 | 910 | 0.62 | 0.8 | 0.8 | 0.44 | 0.66 | 0.73 | 0.74 | 0.59 |
| 27899 | Unlisted procedure, leg or ankle | 166 | 2.69 | 0.85 | 0.92 | 0.92 | 0.79 | 510 | 0.67 | 0.71 | 0.71 | 0.64 | 0.67 | 0.73 | 0.73 | 0.61 |
| 24400 | Osteotomy, humerus, with or without internal fixation | 165 | 0.36 | 0.93 | 0.99 | 0.99 | 0.88 | 910 | 0.66 | 0.9 | 0.9 | 0.42 | 0.75 | 0.75 | 0.75 | 0.76 |
| 27428 | Ligamentous reconstruction (augmentation), knee; intra-articular (open) | 161 | 2.15 | 0.93 | 0.89 | 0.89 | 0.97 | 510 | 0.72 | 0.94 | 0.94 | 0.5 | 0.8 | 0.82 | 0.82 | 0.78 |
| 29325 | Application of hip spica cast; 1 and one-half spica or both legs | 161 | 0.72 | 0.96 | 0.99 | 0.99 | 0.94 | 80 | 0.86 | 0.88 | 0.88 | 0.84 | 0.84 | 0.9 | 0.9 | 0.78 |
| 27305 | Fasciotomy, iliotibial (tenotomy), open | 160 | 0.45 | 0.95 | 0.99 | 0.99 | 0.91 | 510 | 0.62 | 0.96 | 0.97 | 0.28 | 0.79 | 0.79 | 0.79 | 0.78 |
| 29897 | Arthroscopy, ankle (tibiotalar and fibulotalar joints), surgical; debridement, limited | 156 | 0.96 | 0.96 | 0.98 | 0.98 | 0.94 | 510 | 0.66 | 0.95 | 0.96 | 0.35 | 0.83 | 0.88 | 0.88 | 0.77 |
| 29834 | Arthroscopy, elbow, surgical; with removal of loose body or foreign body | 153 | 0.39 | 0.99 | 0.99 | 0.99 | 1 | 910 | 0.73 | 0.97 | 0.97 | 0.48 | 0.92 | 0.87 | 0.87 | 0.97 |
| 20900 | Bone graft, any donor area; minor or small (eg, dowel or button) | 151 | 2.16 | 0.86 | 0.91 | 0.91 | 0.8 | 80 | 0.74 | 0.8 | 0.81 | 0.67 | 0.76 | 0.71 | 0.71 | 0.8 |
| 28300 | Osteotomy; calcaneus (eg, Dwyer or Chambers type procedure), with or without internal fixation | 150 | 0.21 | 0.98 | 0.99 | 0.99 | 0.97 | 510 | 0.5 | 1 | 1 | 0 | 0.86 | 0.86 | 0.86 | 0.87 |
| 27705 | Osteotomy; tibia | 149 | 1.59 | 0.92 | 0.95 | 0.95 | 0.9 | 80 | 0.6 | 0.97 | 0.97 | 0.23 | 0.73 | 0.83 | 0.83 | 0.63 |
| 20205 | Biopsy, muscle; deep | 149 | -0.21 | 1 | 0.99 | 0.99 | 1 | 80 | 0.62 | 1 | 1 | 0.23 | 0.85 | 0.94 | 0.94 | 0.77 |
| 29851 | Arthroscopically aided treatment of intercondylar spine(s) and/or tuberosity fracture(s) of the knee, with or without manipulation; with internal or external fixation (includes arthroscopy) | 148 | -0.14 | 0.99 | 0.99 | 0.99 | 1 | 90 | 0.72 | 0.98 | 0.98 | 0.47 | 0.89 | 0.88 | 0.88 | 0.9 |
| 27691 | Transfer or transplant of single tendon (with muscle redirection or rerouting); deep (eg, anterior tibial or posterior tibial through interosseous space, flexor digitorum longus, flexor hallucis longus, or peroneal tendon to midfoot or hindfoot) | 147 | 0.56 | 0.96 | 0.98 | 0.99 | 0.93 | 510 | 0.58 | 0.99 | 0.99 | 0.17 | 0.84 | 0.86 | 0.86 | 0.83 |
| 29837 | Arthroscopy, elbow, surgical; debridement, limited | 147 | 0.75 | 0.98 | 0.99 | 0.99 | 0.97 | 510 | 0.62 | 0.97 | 0.97 | 0.28 | 0.88 | 0.87 | 0.87 | 0.9 |
| 27425 | Lateral retinacular release, open | 145 | 1.32 | 0.94 | 0.95 | 0.95 | 0.93 | 510 | 0.71 | 0.9 | 0.9 | 0.52 | 0.87 | 0.92 | 0.92 | 0.83 |
| 21899 | Unlisted procedure, neck or thorax | 145 | 1.01 | 0.97 | 0.98 | 0.98 | 0.97 | 510 | 0.72 | 0.99 | 1 | 0.45 | 0.94 | 0.96 | 0.96 | 0.93 |
| 29035 | Application of body cast, shoulder to hips; | 141 | 0.45 | 1 | 0.99 | 0.99 | 1 | 910 | 0.75 | 0.99 | 1 | 0.5 | 0.93 | 0.96 | 0.96 | 0.89 |
| 27502 | Closed treatment of femoral shaft fracture, with manipulation, with or without skin or skeletal traction | 140 | 0.22 | 0.98 | 0.99 | 0.99 | 0.96 | 940 | 0.74 | 0.92 | 0.92 | 0.57 | 0.92 | 0.91 | 0.91 | 0.93 |
| 20926 | Tissue grafts, other (eg, paratenon, fat, dermis) | 137 | 0.98 | 0.96 | 0.99 | 0.99 | 0.93 | 910 | 0.52 | 1 | 1 | 0.04 | 0.78 | 0.81 | 0.81 | 0.74 |
| 24685 | Open treatment of ulnar fracture, proximal end (eg, olecranon or coronoid process[es]), includes internal fixation, when performed | 137 | 0.64 | 0.99 | 0.98 | 0.98 | 1 | 510 | 0.65 | 0.71 | 0.71 | 0.59 | 0.79 | 0.76 | 0.76 | 0.81 |
| 29065 | Application, cast; shoulder to hand (long arm) | 136 | 2.28 | 0.91 | 0.93 | 0.93 | 0.89 | 510 | 0.67 | 0.93 | 0.93 | 0.41 | 0.74 | 0.79 | 0.79 | 0.7 |
| 27690 | Transfer or transplant of single tendon (with muscle redirection or rerouting); superficial (eg, anterior tibial extensors into midfoot) | 133 | 0.5 | 0.98 | 0.99 | 0.99 | 0.96 | 130 | 0.79 | 0.99 | 0.99 | 0.59 | 0.85 | 0.86 | 0.86 | 0.85 |
| 29450 | Application of clubfoot cast with molding or manipulation, long or short leg | 131 | 1.24 | 0.98 | 0.95 | 0.95 | 1 | 90 | 0.54 | 0.99 | 1 | 0.08 | 0.76 | 0.87 | 0.88 | 0.65 |
| 25607 | Open treatment of distal radial extra-articular fracture or epiphyseal separation, with internal fixation | 130 | 1.23 | 0.93 | 0.98 | 0.98 | 0.88 | 510 | 0.67 | 0.77 | 0.77 | 0.58 | 0.76 | 0.78 | 0.78 | 0.73 |
| 27686 | Lengthening or shortening of tendon, leg or ankle; multiple tendons (through same incision), each | 128 | 1.65 | 0.97 | 0.97 | 0.97 | 0.96 | 500 | 0.59 | 0.99 | 0.99 | 0.19 | 0.91 | 0.87 | 0.87 | 0.96 |
| 25390 | Osteoplasty, radius OR ulna; shortening | 127 | -0.08 | 0.99 | 0.99 | 0.99 | 1 | 500 | 0.83 | 0.95 | 0.95 | 0.72 | 0.89 | 0.86 | 0.86 | 0.92 |
